# Feeding behaviour and its determinants among healthy toddlers in an urban city of India

**DOI:** 10.1101/2020.04.16.20067595

**Authors:** Kameswari C Bhairavabhatla, Venkatarao Epari, Sandeep Kumar Panigrahi

**Affiliations:** Bankura Sammilani Medical College and Hospital, Gobindanagar, Bankura, West-Bengal - 722101; Ex-Intern, IMS and SUM Hospital, Siksha ‘O’ Anusandhan deemed to be University, Bhubaneswar, India – 751003; Professor, Community Medicine Department, IMS and SUM Hospital, Siksha ‘O’ Anusandhan deemed to be University, Bhubaneswar, India – 751003; Associate Professor, Community Medicine Department, IMS and SUM Hospital, Siksha ‘O’ Anusandhan deemed to be University, Bhubaneswar, India – 751003

**Keywords:** body mass index, feeding behaviour, food preferences, growth and development, malnutrition, preschool

## Abstract

**Introduction:** Growth and development of a child is largely influenced by feeding behaviour. Feeding problem in infancy and childhood is an important aspect since this may be associated with under-nutrition or even childhood obesity. Social, environmental and emotional factors, type and taste of food, perceptions and practices of parents, etc. may all determine feeding behaviour of child. This study tries to find out the proportion of normal toddlers presenting with feeding problem, factors associated with feeding behaviour and their association with physical growth among these toddlers.

**Methods:** A total of 100 mothers of 1 to 3 year-old children visiting for immunization were interviewed using systematic sampling method. A tool was developed by modifying existing tools are review of literature, was pre-tested and used. Association of various factors with eating behaviour and body mass index (BMI) were studied. Variables measured included perceived feeding problem, anthropometric measures, socio-demographic, emotional, environmental, parental factors, etc. Analysis was done using Chi-square test, t-test or Mann-Whitney U test using IBM SPSS v 20.0.

**Results:** Median age of the children was 19 months. 35% had a feeding problem. None of the socio-demographic variables were found to have any association with feeding problem. All children born preterm had eating problem. Feeding problem got undetected if fed by anyone other than mother. The mean TV viewing time for children with eating problem was significantly more (p = 0.017). Education status of mothers had a positive association with body mass index of the child (p=0.010). Anger, forced feeding and sleepy/drowsiness were reported to decrease feeding while happiness, caressing while feeding, semi-solid and liquid foods, eating with siblings and friends, and hearing short stories were reported to increase.

**Conclusion:** Feeding problem was present in 35% of the toddlers. Preterm birth, long hours of TV viewing during feeding, are associated with eating problem. Mothers are the best persons to identify this problem. Better maternal education decreases the burden of under nourished children though it increases those of overweight and obese.

## 1. Introduction

India is said to possess a double burden of disease – undernutrition and overnutrition, and children in India are the most affected. The burden of underweight children under five years in India has decreased by 7% in a decade time (42.5% in 2005-06 as per National Family Health Survey (NFHS)-3 to 35.7% in 2015-16 as per NFHS-4),[1,2] but still this amounts to one-third of these children, make them prone to illness and death in early life.[2] World Health Organization (WHO) has acknowledged an “obesity epidemic”[3] in the past among adults leading to premature illness and death in later life, that has also started affecting India currently. This is also due to the increase in availability of energy dense foods and sedentary life styles during childhood.[4,5]

Feeding problem in infancy is an important aspect since this may be associated with under-nutrition or even childhood obesity[6]. Feeding problem has got no standard classification or definition system. Feeding problems are also heterogenous in nature, and may consist of problems like food aversion, food selectivity or pickiness, partial to total food refusal, pica, etc.[7] Feeding problem may be seen in as much as 25% of the normally developing children.[8] However, there are no studies in India with a high burden of malnutrition regarding estimates of the same.

Identification of feeding problem in a child during infancy and during childhood phase is a difficult task, particularly in toddlers where every mother believes that their children might be having a feeding problem and all the assessment is based on parent’s perception. Parental assessment tools such as Children’s Eating Behaviour Questionnaire (CEBQ), Screening tool for feeding problems (STEP) and STEP-CHILD though available are not exclusively for toddlers.[6,7,9,10]

A balanced and adequate diet that is determined by feeding behaviour, facilitates physical and mental development of a child. Eating behaviours evolve during the first years of life and evidence indicates that habits acquired in childhood persist through to adulthood.[11] Parents and caregivers play vital role in structuring children’s early dietary experiences. These experiences are linked to children’s eating behaviour and nutritional status.[12] Feeding behaviour are learned and adopted during infancy and early childhood just like other components and once in place are much more difficult to modify. Therefore, adopting healthy behaviours is critical for preschool children before unhealthy practices sneak in.[13] Children usually learn what, when and how much to eat based on the transmission of familial and cultural beliefs, attitudes and practices surrounding food and eating. Parents, particularly mothers are said to be “gate keepers” of children’s eating environment.[12] Their feeding practices strongly influence children’s eating patterns, which are firmly established by five years of age and determine adult eating habits.[14]

Children may also behave differently under a given situation or setting. Social, environmental and emotional factors, type and taste of food, parents’ perceptions and practices, etc. may all determine a child’s feeding behaviour. Thus, it is important to know the factors that improve child feeding and the ones that deprive it. This study has tried to find out the proportion of normal toddlers presenting with feeding problem, association between factors with feeding problem and growth pattern among toddlers, which has never been done in Indian setting.

## 2. Materials & Methods

The study was a hospital based study conducted over a period of six months from April 2016 to October 2016 at the immunization and health guidance clinic of a leading tertiary care hospital in Eastern part of India. The study participants were toddlers (1-3 years), and respondents were their mothers. Toddlers living exclusively in urban area of the city and visiting immunization and child health clinic were included in the study. Children having congenital problems of the gastro-intestinal tract, cerebral palsy or other neuropsychiatric conditions, which would be hindrance from feeding were selectively excluded from the study.

Since this study has never been done among toddlers in India, it was assumed that the prevalence of feeding problem was 50%, and with an absolute precision of 10%, a total sample of 97 would be required to complete the study. A large absolute precision was taken since this was a study never undertaken among toddlers in India, and the prevalence of feeding problems would be enumerated initially for further research. Considering drop outs and non-response rate, an additional 10% of the sample would be required. Thus, a total of 107 samples was considered as the final sample for the study. Systematic sampling method was used in the study.

In every 30 minutes, a mother carrying her child (toddler) to the outpatient for immunization or health guidance, was approached and her informed consent to participate in the ongoing study was taken. On agreeing to participate they were interviewed. Those with medical problems (like heart, renal, congenital malformations, or having undergone any major surgeries) or mothers who were not having sufficient time to interact, were excluded from the study. Similarly, wherever mother of the toddler was not among the caregivers, or had not visited along with, were excluded from the study.

Tool was developed based on available review of literature after modifying validated tools available for data collection. [6,7,9,10] Expert opinion was taken from two paediatricians and two dieticians from the institute before finalizing the tool. The tool was tested before data collection. Mothers are regarded as the primary care takers of children and are the best persons having knowledge regarding their children and their feeding patterns and/or problems. Hence mothers were taken as key informants in this study with regards to feeding behaviour of their child, and based on their perception regarding feeding behaviour of their child, their child was classified as have feeding problem or not. Feeding problem was defined for this study as delays and/or disorders in the development of eating and drinking skills like disordered placement of food in the mouth, difficulty in appropriately manipulating food when it is in the mouth, difficulty chewing, and/or difficulty swallowing. Feeding problems also include deficits in “any aspect of taking nutritional elements that result in under nutrition, poor growth or stressful mealtimes for children and their caregivers”.[15] The factors (emotional state, type and nature of food, and socio-environmental factors) associated with increased or decreased feeding quantity were explored.

Data were collected regarding socio-demographic factors, anthropometric measures, feeding details and feeding behaviour of the child. Socio-demographic data included religion, type of family (nuclear/joint), per capita income, number of family members, mother’s working status and education. Child’s details included age in completed months, time of birth (term/preterm), birth weight, breastfeeding and other feeding habits. Feeding problem (present/absent) as perceived by the mother, was recorded. Effect of emotions, type and nature of food and the environment, on child’s feeding behaviour was elicited on a Likert scale of 0 to 10 [with a score of 5 meant ‘no effect’, towards 0 (zero) had decreasing feeds’ quantity while towards 10 (ten) had increasing feeds’ quantity].

Height and weight of the toddlers up to 2 years were measured using an infantometer. For children 2 to 3 years, weight was measured to the nearest 0.1 kg using a standardized electronic weighing scale with children in light dress and without footwear while height was measured to the nearest cm using a stadiometer. BMI was calculated as weight (in kg) divided by square of height (in metres). Children were grouped as per z-scores based on standard WHO growth charts of BMI for age based on prescribed age and gender. Data were analysed using IBM SPSS v 20.0 licensed to the institute. Continuous data were expressed in terms of mean and standard error (SE) of mean and categorical data as proportions and SE of proportion. Pearson’s chi-square test was used to find associations between categorical variables and t-test / ANOVA for comparing means between groups. However, appropriate non-parametric tests were used wherever data was found to be skewed. A ‘p’ value of < 0.05 was considered as statistically significant in all cases.

Institute Ethics Committee clearance was obtained prior to commencement of the study. Privacy and confidentiality was maintained at all levels, and informed consent was taken prior to the data collection and participation in the study. No monetary benefits or incentives of any kind was provided at any stage for participating in the study.

## 3. Results

The study was conducted among a final target sample of 100 participants after approaching a total of 113 subjects (88.5%). This sample was more than the minimum sample size estimated before onset of the study. Nine were excluded since they had not visited with their mothers. Four participants could not be included in the study for reasons such as no time to participate (2 subjects), not willing to participate (1 subject) and not meeting inclusion criteria (1 subject having nephrotic syndrome). In all cases, the key informants were mother of the toddlers (100 cases, 100%).

The median age of the study group was 19 months (IQR: 16, 24). There were 60 boys (60%) and 40 girls (40%) among the participants. Most children (69%) belonged to joint families. The median per capita income was INR 5,000 per month (IQR: 2,000, 10,000 INR). Most of the mothers (59%) were graduates and above. Majority (84%) of them were however housewives and regular eating habit was present in 87% of mothers (not shown in table). Exclusive breast feeding was found to be given to children for 6 months (Q2: 6 mon; IQR: 4, 6 months) and breast feeding was continued up 17 months of age (Q2: 17 mon; IQR: 12, 22 months). Around 35 children (35%) had perceived feeding problem, as stated by their mothers. None of the socio-demographic factors, including graduation status of either parents, were found to be significantly different in between children having and not having feeding problem (p>0.05) (see table 1). Mid-upper arm circumference, considered as an early marker for identification of severe acute malnutrition (SAM), was also not having any statistical significance in between those with perceived feeding problems and those without. However, overweight or obesity was seen more commonly among toddlers of graduate mothers (47.5% overweight/ obese whereas 14% underweight among graduate mothers) and this association was found to be statistically significant (p=0.010) (not shown in table). The mean television (TV) viewing time for children having feeding problem was 50 minutes/ day, while it was 25 minutes/ day in those not having one (p = 0.017, statistically significant) (Table 1).

**Table 1:**
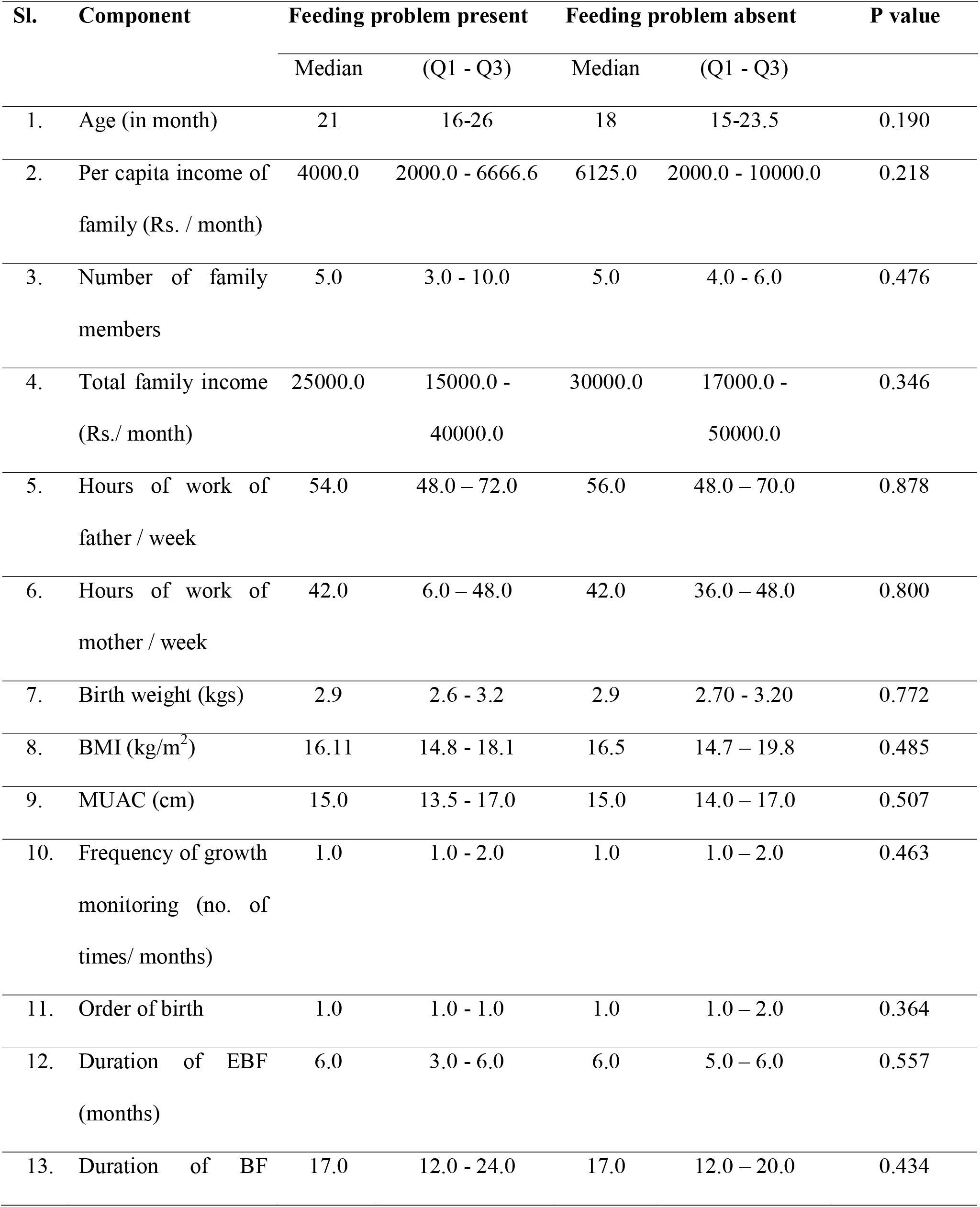
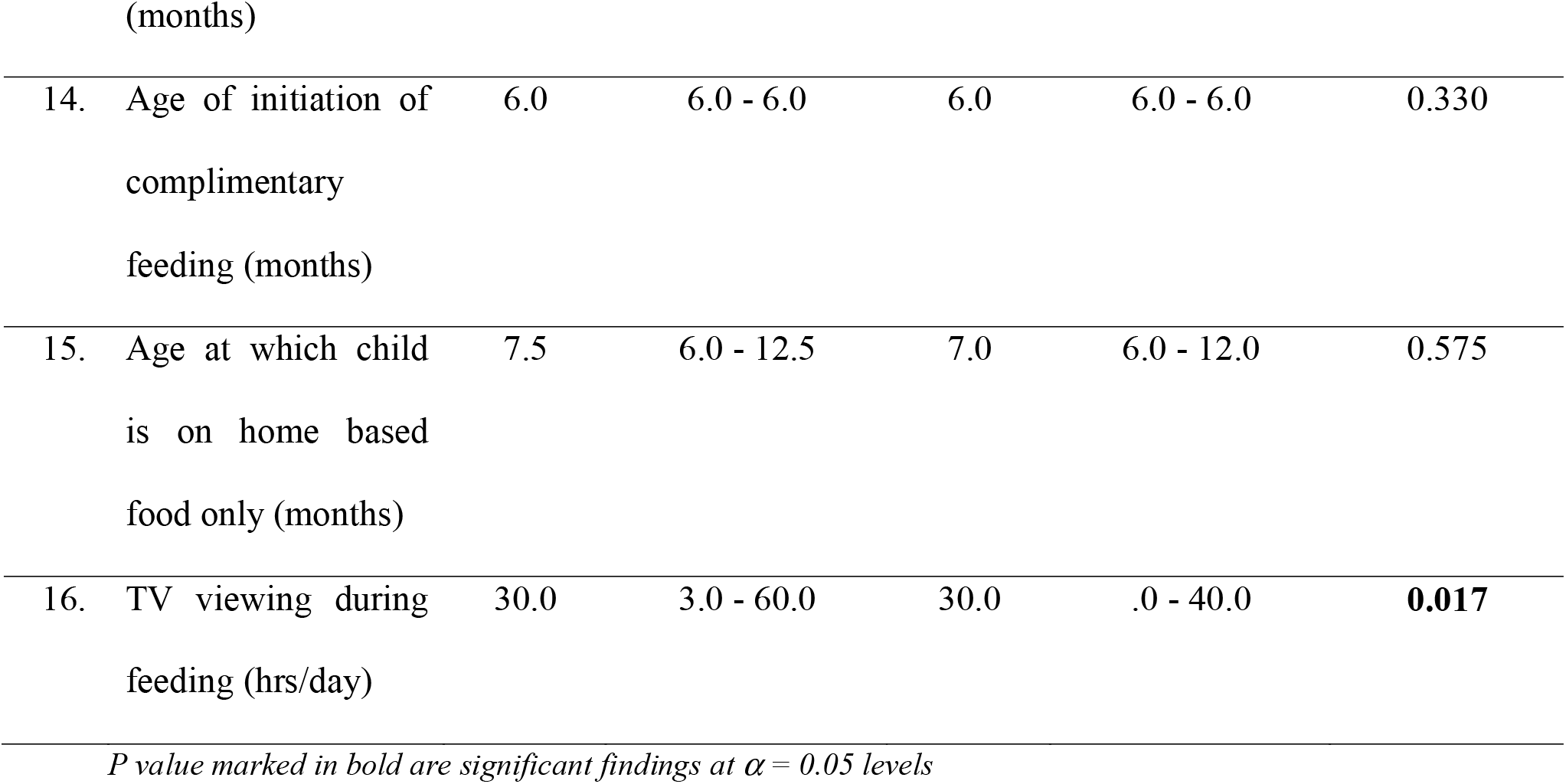
Factors (quantitative variables) determining feeding behaviour of toddlers (n=100)

| Sl. | Component | Feeding problem present |  | Feeding problem absent |  | P value |
| --- | --- | --- | --- | --- | --- | --- |
|  |  | Median | (Q1 - Q3) | Median | (Q1 - Q3) |  |
| 1. | Age (in month) | 21 | 16-26 | 18 | 15-23.5 | 0.190 |
| 2. | Per capita income of family (Rs. / month) | 4000.0 | 2000.0 - 6666.6 | 6125.0 | 2000.0 - 10000.0 | 0.218 |
| 3. | Number of family members | 5.0 | 3.0 - 10.0 | 5.0 | 4.0 - 6.0 | 0.476 |
| 4. | Total family income (Rs./ month) | 25000.0 | 15000.0 - 40000.0 | 30000.0 | 17000.0 - 50000.0 | 0.346 |
| 5. | Hours of work of father / week | 54.0 | 48.0 – 72.0 | 56.0 | 48.0 – 70.0 | 0.878 |
| 6. | Hours of work of mother / week | 42.0 | 6.0 – 48.0 | 42.0 | 36.0 – 48.0 | 0.800 |
| 7. | Birth weight (kgs) | 2.9 | 2.6 - 3.2 | 2.9 | 2.70 - 3.20 | 0.772 |
| 8. | BMI (kg/m<sup>2</sup>) | 16.11 | 14.8 - 18.1 | 16.5 | 14.7 – 19.8 | 0.485 |
| 9. | MUAC (cm) | 15.0 | 13.5 - 17.0 | 15.0 | 14.0 – 17.0 | 0.507 |
| 10. | Frequency of growth monitoring (no. of times/ months) | 1.0 | 1.0 - 2.0 | 1.0 | 1.0 – 2.0 | 0.463 |
| 11. | Order of birth | 1.0 | 1.0 - 1.0 | 1.0 | 1.0 – 2.0 | 0.364 |
| 12. | Duration of EBF (months) | 6.0 | 3.0 - 6.0 | 6.0 | 5.0 – 6.0 | 0.557 |
| 13. | Duration of BF | 17.0 | 12.0 - 24.0 | 17.0 | 12.0 – 20.0 | 0.434 |

Feeding problem was perceived to be present in all the toddlers who had a history of preterm birth (3 cases, 100% of preterm) and it was less in cases who were born at term (32 cases, 33% of 97 term children) and this was found to be statistically significant (p=0.017). There were 93 cases who were found to be fed by mothers and out of these 35 cases (37.6%) reported to have a feeding problem, while none of those fed by caregivers other than mother reported a feeding problem (p = 0.043, statistically significant) (Table 2).

**Table 2:**
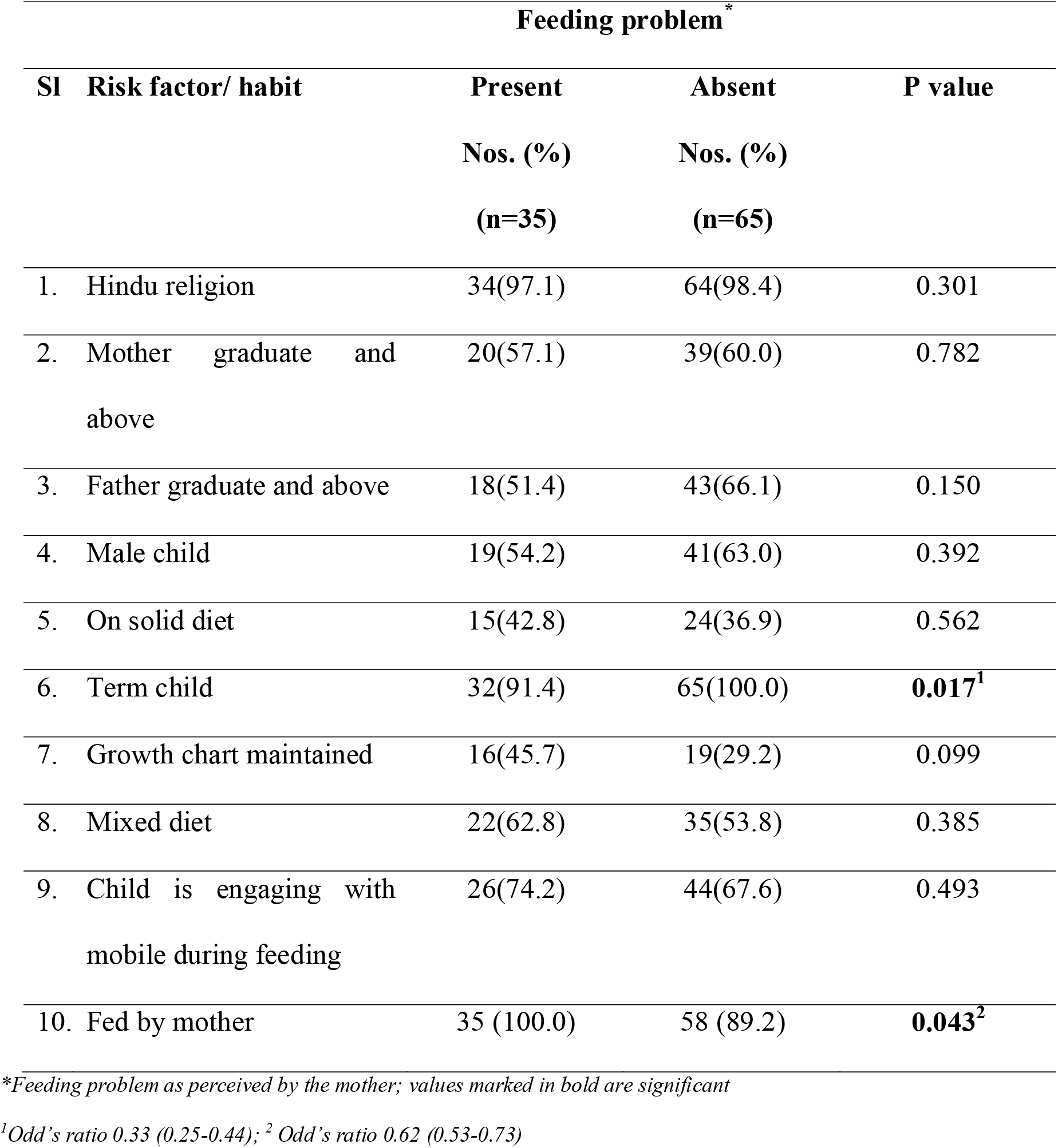
Determinants (categorical variables) of feeding problem as perceived by the mother (n=100)

Among the emotional factors, having anger and being tired were found to decrease the quantity of feeds, and this was found to be statistically significant (p<0.05). Similarly in children having feeding problem, semi-solid foods were found to decrease the quantity of feeds in toddler group (p=0.043). No socio-environmental factors like eating with siblings and friends and hearing short stories were found to have any effect on feeding behaviour of the child (Table 3).

**Table 3:** Association of perceived feeding behaviour of toddlers with the quantity of feeds under a given situation/ circumstance (n=100)

| Factor/ Situation (n)<sup>1</sup> | Quantity of feed |  |  | Feeding Problem<sup>3</sup> | p value |
| --- | --- | --- | --- | --- | --- |
|  | Decreased | No change | Increased |  |  |
|  | (%)<sup>2</sup> | (%)<sup>2</sup> | (%)<sup>2</sup> |  |  |
| EMOTIONAL STATE |  |  |  |  |  |
| Anger (n=98) | 30 (40.0) | 0 (0.0) | 5 (62.5) | 35 (35.7) | **0.003** |
| Upset (n=98) | 22 (45.8) | 8 (23.5) | 5 (31.3) | 35 (35.7) | 0.107 |
| Happy (n=100) | 2 (100.0) | 6 (31.6) | 27 (34.2) | 35 (35.0) | 0.147 |
| Tired (n=100) | 21 (40.4) | 1 (4.8) | 13 (48.1) | 35 (35.0) | **0.004** |
| Frightened (n=97) | 19 (40.4) | 4 (19.0) | 12 (41.4) | 35 (36.1) | 0.185 |
| Without parents (n=97) | 20 (40.0) | 8 (33.3) | 7 (30.4) | 35 (36.1) | 0.694 |
| Forced feeding (n=98) | 26 (35.6) | 2 (20.0) | 6 (40.0) | 34 (34.7) | 0.558 |
| Caressing while feeding (n=100) | 8 (42.1) | 6 (31.6) | 21 (33.9) | 35 (35.0) | 0.758 |
| TYPE AND NATURE OF FOOD |  |  |  |  |  |
| Spicy food (n=84) | 17 (36.2) | 0 (0.0) | 15 (46.9) | 32 (38.1) | 0.123 |
| Sweet food (n=96) | 17 (36.2) | 1 (12.5) | 16 (39.0) | 34 (35.4) | 0.353 |
| Coloured food (n=75) | 5 (25.0) | 6 (28.6) | 17 (50.0) | 28 (37.3) | 0.115 |
| Junk food (n=54) | 7 (28.0) | 1 (100.0) | 16 (57.1) | 24 (44.4) | 0.055 |
| Food from outside (n=70)<sup>1</sup> | 7 (35.0)<sup>2</sup> | 7 (63.6)<sup>2</sup> | 14 (35.9)<sup>2</sup> | 28 (40.0)<sup>2</sup> | 0.218 |
| Solid food (n=96) | 12 (38.7) | 6 (33.3) | 15 (31.9) | 33 (34.4) | 0.822 |
| Semi-solid food (n=100) | 5 (71.4) | 13 (41.9) | 17 (27.4) | 35 (35.0) | **0.043** |
| Liquid food (n=100) | 3 (50.0) | 8 (50.0) | 24 (30.8) | 35 (35.0) | 0.248 |
| SOCIO-ENVIRONMENTAL FACTORS |  |  |  |  |  |

| **Factor/ Situation (n)<sup>1</sup>** | **Quantity of feed** |  |  | **Feeding Problem<sup>3</sup>** | **p value** |
| --- | --- | --- | --- | --- | --- |
|  | **Decreased (%)<sup>2</sup>** | **No change (%)<sup>2</sup>** | **Increased (%)<sup>2</sup>** |  |  |
| Eating with parents (n=97) | 20 (44.4) | 3 (27.3) | 11 (26.8) | 34 (35.1) | 0.197 |
| Eating with siblings (n=77) | 6 (42.9) | 3 (33.3) | 19 (35.2) | 28 (36.4) | 0.851 |
| Eating with friends (n=70) | 8 (50.0) | 4 (36.4) | 15 (34.9) | 27 (38.6) | 0.562 |
| Eating in a gathering (n=75) | 11 (32.4) | 0 (0.0) | 15 (36.6) | 26 (34.7) | 0.701 |
| Eating in a restaurant (n=71) | 13 (38.2) | 2 (40.0) | 11 (34.4) | 26 (36.6) | 0.936 |
| While at someone's home (n=100) | 14 (36.8) | 9 (27.3) | 12 (41.4) | 35 (35.0) | 0.486 |
| While hearing short stories (n=99) | 3 (42.9) | 8 (33.3) | 23 (33.8) | 34 (34.3) | 0.885 |
| TV viewing (n=99) | 3 (37.5) | 7 (21.2) | 25 (43.1) | 35 (35.4) | 0.109 |
| AC room (n=91) | 7 (33.3) | 13 (39.4) | 13 (35.1) | 33 (36.3) | 0.888 |
| If sleepy or drowsy (n=100) | 23 (35.9) | 0 (0.0) | 12 (42.9) | 35 (35.0) | 0.078 |

<sup>1</sup>though all respondents answered the questions (n=100), situation was not applicable to some of them. Hence n is determined individually for a given situation, eg. n=70 for food from outside means there are 30 toddlers who do not eat food from outside
<sup>2</sup>Percentage is out of total of same category (i.e. with and without feeding problem), eg. If there are 7 toddlers with feeding problem whose quantity of feed decrease when taking food from outside, there are 13 without feeding problem whose quantity of feed decrease when taking food from outside, thus amounting to 7/20 or 35%
<sup>3</sup>Feeding problem is the total toddlers in a given situation who were perceived by mother to have feeding problem, percentage is out of total responded for that given situation

## 4. Discussion

Eating behaviour of a child (external eating, restriction and emotional eating) is known to contribute to the difference in their weight and their growth. Appetite control, determined by interaction between homeostatic and hedonic mechanisms, that develops during the toddler age also contributes to the child’s eating behaviour. Hedonic mechanisms involved in child’s eating behaviour development are usually modifiable and depend on the role a family plays in developing a child’s eating behaviour.[6]

Feeding behaviour of a child is related to the smell and taste perception, a part of the hedonic mechanism.[6] Taste perception develops as early as the fetal period with an exposure to the amniotic fluid (taste similar to garlic, cumin and curry). Later on exposure to mother’s milk also sets a platform for taste perception to certain other categories like garlic, alcohol and vanilla.[6] Bottle-fed infants lose the ability to self-regulate appetite and thus fail to decide the timing and amount of intake.[6]

They are not independent and could interact with each other. The use of the CEBQ allows the identification of children with problematic eating behaviours, regardless of its aetiology. It is a valid method, largely applied in different population settings. The reduced ability of children to respond to internal signals of hunger and satiety can be imposed by parents. Parental child-feeding practices could influence a child’s weight, but they could also be a reaction to the child’s weight – this bidirectional effect should be considered in future studies. Given the close association between appetite-related eating behaviours and weight status, future studies on the determinants of eating behaviours may favour the development of useful tools for prevention, support and treatment of childhood obesity. [6]

Review conducted by Rossi et.al. has shown that family played the key role in forming healthy eating habits, since modifying eating habits during adulthood is difficult once it is established. School, social network, cultural and socioeconomic conditions are potentially modifiable determinants in the process of building the child’s eating habits as in the case of an adult.[16]

In this study, an association was seen in this study between preterm birth and feeding problem. Infants born preterm and/or with a birth weight below 10^th^ percentile for gestational age were found to be at greater risk for developing feeding disorders, even in some studies.[17] It is a well-known fact that preterm babies have growth and development related problems, nutrition being one of them. This study shows results consistent with previous studies.

Feeding problem was found to have significant association with cases being fed by mother, which may seem to be contradicting. This finding probably reflects that mothers who are feeding their children regularly are able to identify feeding problem in their children, and not that children feed poorly when fed by mothers. In present day scenario, where number of working mothers is increasing in our society, more and more children are taken care by care takers at home or at crèches. The diet of such toddlers may get neglected and their feeding problems may thus go unidentified. This has not been demonstrated in any studies around the world, though concepts like Integrated Management of Childhood Illness (IMCI) believes that disease identification in a child can be known better by asking the mother.[18]

Children having feeding problem were found to have significantly more TV viewing time as compared to those who are not. Research shows positive association between TV viewing time and high-energy food consumption and negative associations with vegetable consumption. There is documented relationship between TV viewing, exposure to food advertising, increased requests for advertised foods and preferences for the advertised items.[19]

In a study conducted by Mazur et.al. with an objective as to what extent the psychosocial factors were associated with feeding behaviour in case of adolescents, it was seen that negative behaviours like dietary restrictions, uncontrolled eating and emotional eating possibly had strong association with psychosocial factors in adolescence. Emotional eating was strongly associated with feeding behaviour weakest was with dietary restrictions.[20]

In a review conducted by Scaglioni et.al., so as to understand the details of eating behaviour and devise preventive measures to manage the nutrition of the children, many important factors were observed. The family system surrounding a child’s domestic life had an active role in establishing and promoting persistent behaviour of a child. Experiences in early part of life with various tastes and flavours were seen to promote healthy eating in the future life. Food habits of the parents and feeding strategies were the most important determinants of a child’s eating behaviour and food choices. It was highlighted that parents should expose their offspring to a range of good food choices while acting as positive role models. Whenever prevention programmes were planned they should take into account the socioeconomic aspects and education.[21]

There are multiple factors in the parent and the family which influence the children, and this is always bidirectional. It is known that family environment influences children intake (eating styles, diet composition, availability of energy dense foods at home, eating at home than at restaurants, etc.). Currently parental control like pressurizing children to eat healthy food and diet restriction to avoid unhealthy foods are involved in the eating behaviour of their child.[11]

The cultural context of India differs variedly and it is known that there is gross difference in socio-environmental conditions of people living in rural and urban areas. The study was done in an urban setting and in eastern part of India. Hence this may not be generalizable to entire community, nor even to the entire country. Further research may be needed with a large sample and covering urban and rural population. Longitudinal studies and interventional models may show better results in determining feeding behaviour among toddlers.

## 5. Conclusion

Children born preterm have more eating problems as compared to those born at term. Mothers should personally take care of their children, especially their feeding as they are the best persons who identify eating problems among children. Eating problems of children who are fed by care-takers might be overlooked and neglected. Longer periods of TV viewing during feeding results in eating problems in children, which could be due to the impact of advertised food on TV. Better maternal education decreases chances of their children being under nourished though it may increase overweight and obesity. Parents should take adequate care to avoid situations having negative impact on child feeding such as anger and forced feeding whereas factors which increase socialization like feeding with friends and family might have positive impact on feeding. At the same time, parents should try to promote such environments, parenting styles and food items which are better accepted by children and improve their nutrition.

## 6. Implications for research and practice

Further evidence generation can be done through high order studies such as Cohort studies. The findings from this study can be tested also through well-designed clinical trials, and the findings can be utilized by paediatricians to improve feeding behaviour and foster changes in toddlers visiting their clinic.

## Data Availability

Data will be made available on request to the corresponding author.

## Acknowledgement

The authors acknowledge Indian Council of Medical Research for providing scholarship to the student for conducting this study as a part of STS Project (ID 2016-00314). The authors also acknowledge the review and feedback for the draft manuscript provided by Dr. Jyotiranjan Sahoo, Department of Community Medicine, IMS and SUM Hospital, Bhubaneswar.

